# COVID-19 detection from chest X-Ray images using Deep Learning and Convolutional Neural Networks

**DOI:** 10.1101/2020.05.22.20110817

**Authors:** Antonios Makris, Ioannis Kontopoulos, Konstantinos Tserpes

## Abstract

The COVID-19 pandemic in 2020 has highlighted the need to pull all available resources towards the mitigation of the devastating effects of such “Black Swan” events. Towards that end, we investigated the option to employ technology in order to assist the diagnosis of patients infected by the virus. As such, several state-of-the-art pre-trained convolutional neural networks were evaluated as of their ability to detect infected patients from chest X-Ray images. A dataset was created as a mix of publicly available X-ray images from patients with confirmed COVID-19 disease, common bacterial pneumonia and healthy individuals. To mitigate the small number of samples, we employed transfer learning, which transfers knowledge extracted by pre-trained models to the model to be trained. The experimental results demonstrate that the classification performance can reach an accuracy of 95% for the best two models.

## 1 Introduction

The year 2020 has been marked by the pandemic disease caused by a type of the corona virus family (CoV), called COVID-19 or SARS-CoV-2, which has led to over four million infections and more than 290,000 deaths worldwide. COVID-19 is a Severe Acute Respiratory Syndrome (SARS) that was firstly identified in Wuhan, China in December 2019 and has rapidly spread globally in a few months, making it a highly contagious virus. The virus is characterized by symptoms that mostly relate to the respiratory system and include shortness of breath, loss of smell and taste, cough and fever, a range of symptoms that is shared among other types of viruses such as the common cold.

Unlike other viruses, COVID-19 has a long incubation period, ranging from 3 days to 13 days, although on average, the time from exposure to symptom onset is approximately five to six days. The long incubation period makes COVID-19 more contagious since people carrying the virus will most probably keep interacting with other people until they realize they have the virus, leading to more infections. Furthermore, it has been reported that several patients carrying the disease might not show any symptoms at all (asymptomatic patients). The combination of both the long incubation period and the asymptomatic cases, makes the corona virus even harder to detect, trace and contain, which explains its rapid spread.

Several studies have been conducted since the beginning of the year 2020 that try to develop a methodology to identify patients carrying the disease [1, 2, 3, 4]. The majority of these studies in the field of computer science, employ convolutional neural networks (CNNs) in order to classify images of CT scans or X-Rays of the chest as normal or not, in an attempt to identify possible cases of the corona virus. The widespread usage of CNNs for image classification tasks is due to the fact that they have demonstrated a high-accuracy performance in the fields of image recognition and object detection [5]. Over the years, CNNs became more complex, from the first CNN, LeNet-5 [6] which had 5 layers to the deeper architecture of ResNet-50 [7] which had 152 layers. Their success lies in the fact that they are able to capture hidden features of the images, through their numerous hidden layers.

In this research work the effectiveness of several state-of-the-art pre-trained convolutional neural networks was evaluated regarding the automatic detection of COVID-19 disease from chest X-Ray images. A collection of 336 X-Ray scans in total from patients with COVID-19 disease, bacterial pneumonia and normal incidents is processed and utilized to train and test the CNNs. Due to the limited available data related to COVID-19, the transfer learning strategy is employed. The main difference between our work and the previous studies is that this study incorporates a large number of CNN architectures in an attempt to not only distinguish X-Rays between COVID-19 patients and people without the disease, but to also discriminate pneumonia patients from patients with the corona virus, acting as a classifier of respiratory diseases.

The rest of the paper is structured as follows. Section 2 presents a two-fold literature review: i) the usage of deep learning for image classification and ii) the usage of deep learning for the detection of COVID-19. Section 3 describes the methodology employed towards the identification of the corona virus through X-Ray scans, while Section 4 presents the research findings and the experimental results. Finally, Section 5 concludes the merits of our work and presents roadmaps for future research.

## 2 Related Work

### 2.1 Deep learning approaches for image classification and object detection

Numerous studies have used Convolutional Neural Networks (CNNs) for the problem of image classification in the literature, most of which create different architectures for the neural networks. Deep convolutional neural networks are one of the powerful deep learning architectures and have been widely applied in a broad range of machine learning tasks. According to [8] CNNs are able to handle four different manners: training the weights from scratch in the presence of a very large available dataset, fine-tuning the weights of an existing pre-trained CNN with smaller datasets, unsupervised pre-training for weights initialization before putting inputs into CNN models and pre-training CNN as a feature extractor. The first CNN to create a standard “architectural template” was the LeNet-5 [6], which uses two convolutional layers and three fully-connected ones. Ever since, more architectures followed that used the same idea of adding more convolutions and pooling layers, ending with one or more fully-connected ones. Following the footsteps of the previous CNN, AlexNet [9] added three more convolutional layers, making it the deepest neural network of its time. Moreover, AlexNet was the first CNN architecture that implemented Rectified Linear Units (ReLUs) as an activation function. Before making more variations on the architectures, researchers continued using more layers and creating deeper networks and as a result, VGG-16 [10] was emerged. VGG-16 used 13 convolutional layers and 3 fully connected ones, keeping the ReLUs from AlexNet as an activation function. VGG-19, a successor of the previous network, simply added more layers.

The years that followed, researchers, apart from making the networks deeper, added more complexity by introducing several techniques inside the layers of the networks. Inception-v1 [11] besides the fact that it uses 22 layers in total, it also uses a “network inside a network” approach by using “Inception” modules. The main concept of these modules was to use parallel towers of convolutions with different filters, each filter capturing different features, and then cluster these features together. The idea was motivated by Arora et al. [12], which suggested an architecture that analyzes the correlation statistics of the last layer and clusters them into groups of high-correlation units. Sharing a similar architecture, Inception-v3 [13], a successor of the previous network, was among the first networks to use batch normalization to the layers. Inception-v4 [14], the latest successor of the two previous networks, added more Inception modules and made some modifications to improve the training speed. The same authors of the previous networks introduced a family of a new architecture, called Inception-ResNet-v2 [15], in which they converted the Inception modules to Residual Inception blocks, created a new type of Inception modules and added more of these to the network, making it even deeper. ResNet-50 [7] was also among the first networks to use batch normalization. Moreover, it had an even deeper architecture (152 layers) and it used skip connections or residuals. Xception [16] replaced the Inception modules with depthwise separable convolutions. This means that it performed 1 × 1 convolutions to every channel, and then performed a 3 × 3 convolution to each output. Similarly to Xception, MobileNetV2 [17] uses depthwise separable convolutions, which reduce the complexity and size of the network. Furthermore, a module with inverted residual structure is introduced and non-linearities in narrow layers are removed. The characteristics of this network introduced a state-of-the-art image classifier suitable for mobile devices. Finally, the “battle” for a better network architecture continued and resulted in several other CNNs, each one introducing a different modification, such as DenseNet [18], NASNet [19] and ResNet152V2 [20].

### 2.2 Deep learning approaches for COVID-19 detection based on image classification

Various research studies already exist for COVID-19 detection. For the most part, deep learning techniques are employed on chest radiography images with a view to detect infected patients and the results have been shown to be quite promising in terms of accuracy. In [21] a deep convolutional neural network able to predict the coronavirus disease from chest X-ray (CXR) images is presented. The proposed CNN is based on pre-trained transfer models (ResNet50, InceptionV3 and Inception-ResNetV2), in order to obtain high prediction accuracy from a small sample of X-ray images. The images are classified into two classes, normal and COVID-19. Furthermore, to overcome the insufficient data and training time, a transfer learning technique is applied by employing the ImageNet dataset. The results showed the superiority of ResNet50 model in terms of accuracy in both training and testing stage. Abbas et al [22] presented a novel CNN architecture based on transfer learning and class decomposition in order to improve the performance of pre-trained models on the classification of X-ray images. The proposed architecture is called DeTraC and consist of three phases. In the first phase an ImageNet pre-trained CNN is employed for local feature extraction. In the second phase a stochastic gradient descent optimisation method is applied for training and finally the class-composition layer is adapted for the final classification of the images using error-correction criteria applied to a softmax layer. The ResNet18 pre-trained ImageNet network is used and the results showed an accuracy of 95.12% on CXR images. Zhang et al [23] presented a new deep anomaly detection model for fast, reliable screening of COVID-19 based on CXR images. The proposed model consist of three components namely a backbone network, a classification head and an anomaly detection head. The backbone network extract the high-level features of images, which are then used as input into the classification and anomaly detection head. The classification head is used for image classification and consist of a new classification convolutional layer which contains a hidden layer of 100-neurons, an one-neuron output layer, and the “sigmoid” activation function. The anomaly detection head has the same architecture as the classification but generates the scalar anomaly scores which in turn detects anomaly images (COVID-19 cases). The proposed model achieved to reduce the false positive rate. More specifically, the results demonstrated a sensitivity and specificity of 96.00% and 70.65% respectively. In [24] a deep convolutional neural network called COVID-Net is presented which is able to detect COVID-19 cases from CXR images. The network design is consist of two stages, a human-machine collaborative design strategy and a machine-driven design exploration stage and the architecture utilizes a lightweight residual projection-expansion-projection-extension (PEPX) design pattern. Furthermore, an explainability-driven audit is performed for decisions validation. The results showed a high sensitivity (87.1%) and a precision of 96.4% for COVID-19 cases. Another work [4] presents a CNN framework for COVID-19 detection from other pneumonia cases. The framework called COVID-ResNet and utilizes a three step technique to fine-tune a pre-trained ResNet-50 architecture in order to improve performance and reduce training time. Progressive resizing of input images (28×128×3-stage 1, 224×224×3-stage 2, 229×229×3-stage 3) and fine-tuning of network at each stage manages to achieve a better generalization and an increased overall performance (96.23% accuracy). Hemdan et al [25] presented a framework consist of seven deep learning image classifiers called COVIDX-Net with a view of classifying COVID-19 disease from CXR images. As the results showed, the best performance achieved for the VGG19 and DenseNet201 classifiers with an accuracy of 90%. In [26] the authors investigated how Monte-Carlo Dropweights Bayesian convolutional neural networks can estimate uncertainty in deep learning in order to improve the performance of human-machine decisions. Bayesian Deep Learning classifier has been trained using transfer learning on a pre-trained ResNet50V2 model using COVID-19 X-Ray images to estimate model uncertainty. The results demonstrated a strong correlation between estimated uncertainty in prediction and classification accuracy, thus enabling false predictions identification. Finally, Apostolopoulos et al [1] evaluated the performance of five pre-trained CNN networks regarding the detection of COVID-19 from CXR. The results showed that VGG19 and MobileNetv2 achieved the higher accuracy, 93.48% and 92.85% respectively.

## 3 Methodology

### 3.1 Dataset Description

The dataset used in this study contains chest X-Ray images from patients with confirmed COVID-19 disease, common bacterial pneumonia and normal incidents (no infections) and is a combination of two different publicly available datasets. More specifically, COVID-19 cases have been obtained from Dr. Joseph Cohen’s Github repository [27] and consist of 112 Posterior-Anterior (PA) X-ray images of lungs. In general, this repository contains chest X-ray / CT images of patients with acute respiratory distress syndrome (ARDS), COVID-19, Middle East respiratory syndrome (MERS), pneumonia and severe acute respiratory syndrome (SARS). In addition, 112 normal and 112 pneumonia (bacterial) chest X-Ray images were selected from Kaggle’s repository^2^. In summary, the dataset used for this work is evenly distributed regarding the number of cases and consist of 3 classes (covid, pneumonia and normal) and it is publicly available in^3^. There are some limitations that are worth mentioning. Firstly, confirmed COVID-19 samples exist already is very small compared to pneumonia or normal cases. At this time, there is not a larger and reliable sample available. The same number of samples was selected for each class for the sake of uniformity. Furthermore, to the best of our knowledge the pneumonia samples are older recorded samples and do not represent pneumonia images from patients with suspected coronavirus symptoms, while the clinical conditions are missing. Finally, the normal class represents individuals that are not classified as COVID-19 or pneumonia cases. We do not imply that a “normal” patient based on the CXR image does not have any emerging disease.

### 3.2 Data Augmentation

Data augmentation is a commonly used process in deep learning which increases the number of the available samples. In this work, due to the lack of a larger number of available samples, data augmentation with multiple pre-processing techniques was performed, leveraging Keras *ImageDataGenerator* during training. The transformations that employed include random rotation of the images (maximum rotation angle was 30 degrees), horizontal flips, shearing, zooming, cropping and small random noise perturbation. Data augmentation improves the generalization and enhance the learning capability of the model. Furthermore it is another efficient way to prevent model overfitting by increasing the amount of training data using information only in training [28].

### 3.3 Performance Metrics

The performance metrics adopted are:

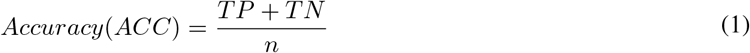

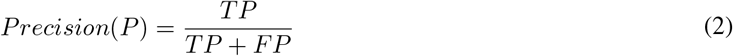

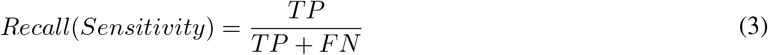

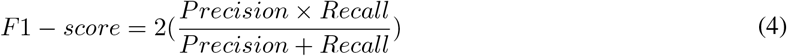

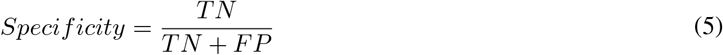

where TP, TN, FP, FN refer to the true positive, true negative, false positive and false negative samples for each class (covid, pneumonia, normal). Then, the macro-average results were computed and used to present the classification performance achieved by the networks.

*Accuracy* is a commonly used classification metric and indicates how well a classification algorithm can discriminate the classes in the test set. As shown in Eq 1, the accuracy can be defined as the proportion of the predicted correct labels to the total number (predicted and actual) of labels. In this study, accuracy refers to the overall accuracy of the model in distinguishing the three classes (covid, pneumonia, normal). *Precision* (Eq 2) is the proportion of predicted correct labels to the total number of actual labels while *Recall* (Eq 3) is the proportion of predicted correct labels to the total number of predicted labels. Recall is often referred as sensitivity (also called true positive rate). Furthermore, *F1 – score* (Eq 4) refers to the harmonic mean of Precision and Recall while *Specificity* (also called true negative rate) measures the proportion of actual negatives that are correctly identified as such (Eq 5).

### 3.4 Transfer learning with CNNs: fine-tuning

Deep learning models require a large amount of data in order to perform accurate feature extraction and classification. Regarding medical data analysis, especially if the disease is at an early stage such as in COVID-19, one major drawback is that the data analyzed were relatively limited. In order to overcome this limitation, transfer learning was adopted. Transfer learning method achieves data training with fewer samples as the retention of the knowledge extracted by a pre-trained model is then transferred to the model to be trained. A pre-trained model is a network that was previously trained on a large dataset, typically on a large-scale image-classification task. The intuition behind transfer learning for image classification is that if a model is trained on a general large dataset, this model will effectively serve in turn as a generic model. The learned features can be used to solve a different but related task involving new data, which usually are of a smaller population to train a CNN from scratch [29]. Thus the need of training from scratch a large model on a large dataset is eliminated.

In general, there are two types of transfer learning in the context of deep learning: a) feature extraction [30] and b) fine-tuning [31, 32], In feature extraction a new classifier will be trained from scratch on top of the pre-trained model. The representations learned from the pre-trained model which treated as an arbitrary feature extractor are employed in order to extract meaningful features from new samples. The base convolutional network already contains generically useful features for classification, thus there is no need for retraining the entire model. On the other hand, for an increased performance, in fine-tuning the weights of the top layers of the pre-trained model are “fine-tuned” along with the newly-added classifier layers. Thus, the weights are tuned from generic feature maps to features associated specifically with the provided dataset. The aim of fine-tuning is to adapt specialized features to a given task rather than overwrite the generic learning. Fine-tuned learning experiments are much faster and more accurate compared to models trained from scratch [33].

In this work, the CNN models were fine-tuned to identify and classify the different classes (covid, pneumonia, normal). The weights used by all CNNs are pre-trained on the ImageNet dataset [34], ImageNet is an image database which contains about 14 million images belonging to more than 20.000 categories created for image recognition competitions. Figure 1 illustrates an example of the fine-tuning process on the VGG16 network architecture. The network is instantiated with weights pre-trained on ImageNet. On the top of the figure the layers of the VGG16 network are showed. As stated in 2.1, VGG16 contains 13 convolutional (*CONV*) and 3 fully-connected (*FC*) layers. The final set of layers which contain the *FC* layers along with the *softmax* activation function is called “head”. Afterwards, the *FC* layers are excluded and the final *POOL* layer is treated as a feature extractor as depicted in the middle of the figure. Finally, a new *FC* head layer is randomly initialized and placed on top of the original architecture (bottom of the figure). Is is worth mentioning, that the body of the network, i.e. the *CONV* layers have been “repressed” such that only the *FC* head layer is trained. The reason for this behaviour is that the *CONV* layers have already learned discriminative filters while *FC* head layer is randomly initialized from scratch and random values are able to destroy the learned features.

**Figure 1:**
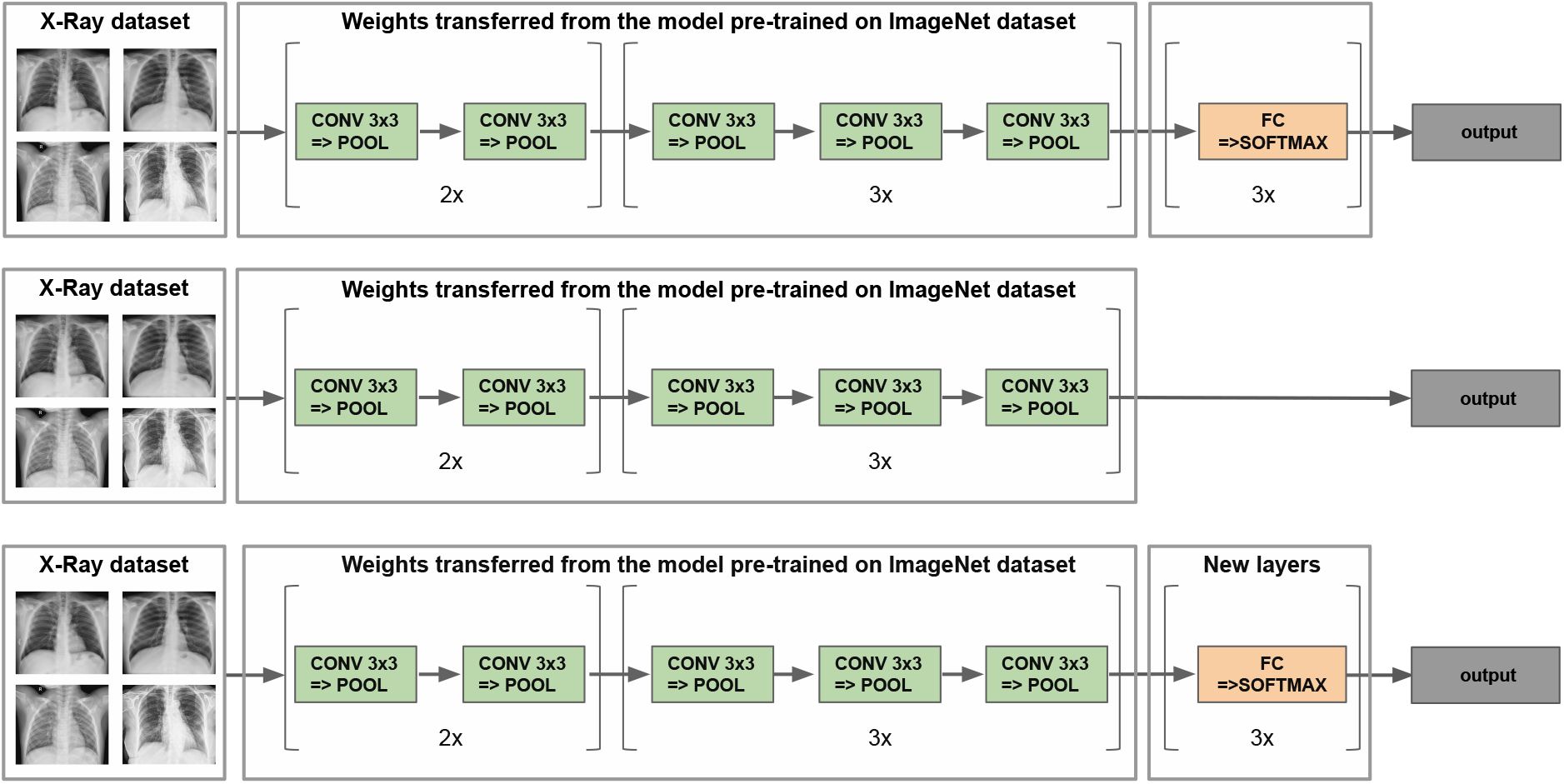
Fine-tuning on the VGG16 network architecture.

## 4 Experimental evaluation

In this research work the effectiveness of several state-of-the-art pre-trained convolutional neural networks was evaluated regarding the detection of COVID-19 disease from chest X-Ray images. More specific, a pool of existing deep learning classifiers were employed namely, VGG16, VGG19, MobileNet V2, Inception V3, Xception, InceptionResNet V2, DenseNet201, ResNet152 V2 and NASNetLarge.

### 4.1 Experimental Setup

To train the proposed deep transfer learning models, the python programming language was used including the Keras package and a TensorFlow backend. Keras is a simple to use neural network library built on top of Theano or TensorFlow [35]. Keras provides most of the building blocks needed to build reasonably sophisticated deep learning models. This framework was used along with the set of weights learned on ImageNet.

The underlying computing infrastructure that has been used for the execution of the CNNs has been a commodity machine with the following configuration: Ubuntu 18.04 LTS 64-bit; Intel Core i7-8550U CPU @ 1.80GHz × 8; and 16 GiB RAM.

### 4.2 Parameters tuning

All the examined CNNs share some common hyper-parameters. Specifically, all images were scaled to a fixed size of 224 × 224 pixels. The dataset used was randomly split into 80% and 20% for training and testing respectively and the training was conducted for 35 epochs to avoid overfitting for all pre-trained models with a learning rate of 1*e* – 3 and a batch size of 8. CNNs were compiled utilizing the optimization method called Adam [36] and all the convolutional layers are activated by the Rectified Linear Unit (ReLU) [37]. Furthermore, a Dropout layer [38] of 0.5 is applied which means that 50% of neurons will randomly set to zero during each training epoch thus avoiding overfitting on the training dataset. Dropout is a form of regularization that forces the weights in the network to receive only small values making the distribution of weight values more regular. As a result this technique can reduce overfitting on small training examples [39]. Since the problem consists of 3 classes the “categorical_crossentropy” is employed as loss function as shown in Eq 6, where *p_model_* [*y_i_* ∈ *C_yi_*] is the probability predicted by the model for the *i^th^* observation to belong to the *c^th^* category. “Categorical crossentropy” compares the distribution of the predictions with the true distribution. True class is represented as a one-hot encoded vector, and the closer the model’s outputs are to that vector, the lower the loss.

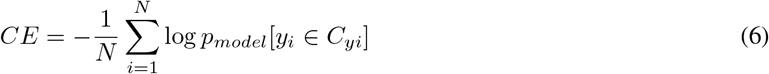

### 4.3 Results & Discussion

In this section the classification performance for each CNN is presented. In order to evaluate the results, the following metrics were adopted for each class (covid, pneumonia, normal): precision, recall (sensitivity), F1-Score, specificity and the overall accuracy of the model as illustrated in Table 1. The results suggest that the VGG16 and the VGG19 achieve the best classification accuracy of 95%. NASNetLarge model showed a moderate accuracy of 81%. The other models did not surpass 80% of accuracy with MobileNetV2 and DenseNet201 presenting the lowest results with 40% and 38% accuracy respectively. Furthrmore, the confusion matrices of the best two models (VGG16, VGG19), the moderate model (NASNetLarge) and the worst models (MobileNetV2 and DenseNet201) are presented in Figure 4.

**Table 1:**
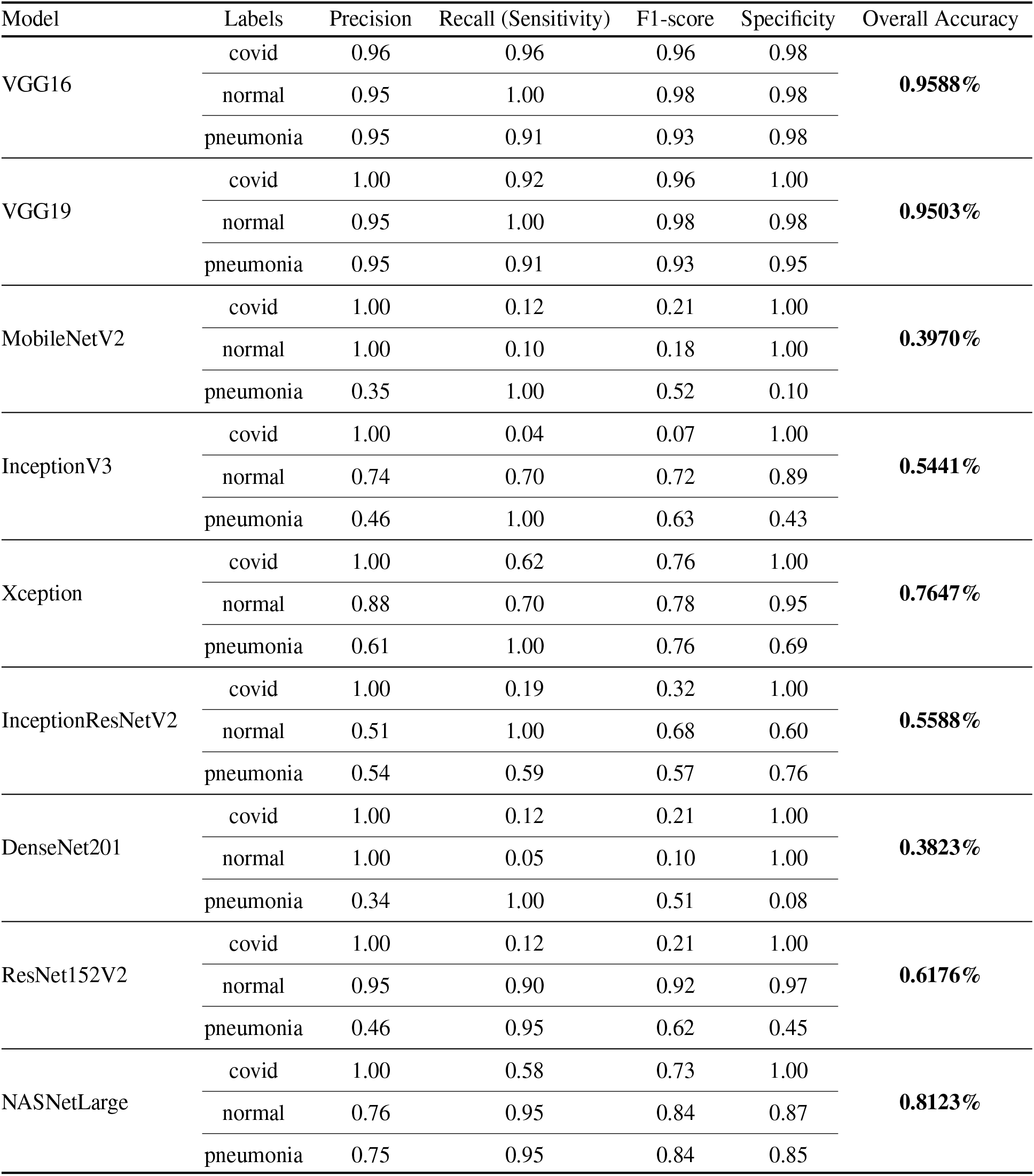
Classification performance obtained from different pre-trained CNN models.

| Model | Labels | Precision | Recall (Sensitivity) | F1-score | Specificity | Overall Accuracy |
| --- | --- | --- | --- | --- | --- | --- |
| VGG16 | covid | 0.96 | 0.96 | 0.96 | 0.98 | **0.9588%** |
|  | normal | 0.95 | 1.00 | 0.98 | 0.98 |  |
|  | pneumonia | 0.95 | 0.91 | 0.93 | 0.98 |  |
| VGG19 | covid | 1.00 | 0.92 | 0.96 | 1.00 | **0.9503%** |
|  | normal | 0.95 | 1.00 | 0.98 | 0.98 |  |
|  | pneumonia | 0.95 | 0.91 | 0.93 | 0.95 |  |
| MobileNetV2 | covid | 1.00 | 0.12 | 0.21 | 1.00 | **0.3970%** |
|  | normal | 1.00 | 0.10 | 0.18 | 1.00 |  |
|  | pneumonia | 0.35 | 1.00 | 0.52 | 0.10 |  |
| InceptionV3 | covid | 1.00 | 0.04 | 0.07 | 1.00 | **0.5441%** |
|  | normal | 0.74 | 0.70 | 0.72 | 0.89 |  |
|  | pneumonia | 0.46 | 1.00 | 0.63 | 0.43 |  |
| Xception | covid | 1.00 | 0.62 | 0.76 | 1.00 | **0.7647%** |
|  | normal | 0.88 | 0.70 | 0.78 | 0.95 |  |
|  | pneumonia | 0.61 | 1.00 | 0.76 | 0.69 |  |
| InceptionResNetV2 | covid | 1.00 | 0.19 | 0.32 | 1.00 | **0.5588%** |
|  | normal | 0.51 | 1.00 | 0.68 | 0.60 |  |
|  | pneumonia | 0.54 | 0.59 | 0.57 | 0.76 |  |
| DenseNet201 | covid | 1.00 | 0.12 | 0.21 | 1.00 | **0.3823%** |
|  | normal | 1.00 | 0.05 | 0.10 | 1.00 |  |
|  | pneumonia | 0.34 | 1.00 | 0.51 | 0.08 |  |
| ResNet152V2 | covid | 1.00 | 0.12 | 0.21 | 1.00 | **0.6176%** |
|  | normal | 0.95 | 0.90 | 0.92 | 0.97 |  |
|  | pneumonia | 0.46 | 0.95 | 0.62 | 0.45 |  |
| NASNetLarge | covid | 1.00 | 0.58 | 0.73 | 1.00 | **0.8123%** |
|  | normal | 0.76 | 0.95 | 0.84 | 0.87 |  |
|  | pneumonia | 0.75 | 0.95 | 0.84 | 0.85 |  |

A sensitivity of 96% and 92% for the covid class can be observed for VGG16 and VGG19 models respectively. This is critical as the model should be able to detect all positive COVID-19 cases to reduce the virus spread to the community. In other words, confirmed positive COVID-19 patients would be accurately identified as “COVID-19 positive” 96% and 92% of the time by employing VGG16 and VGG19 models respectively. Furthermore, the aforementioned models show a high precision value of 96% and 100% for covid class respectively. This implies that for VGG19 there were no classes incorrectly classified as covid from another classes while for VGG16 only one covid case was incorrectly classified as pneumonia as showed in Figures 4(b), 4(a). Another important aspect of the results is the high values associated with specificity. Specifically, the specificity for the covid class is 98% and 100% for VGG16 and VGG19 respectively. This practically means that confirmed negative patients to COVID-19 would be accurately identified as “COVID-19 negative” 98% and 100% of the time using VGG16 and VGG19 models respectively. A similar trend can be depicted in terms of F1-score. Also, one of the very encouraging results is the ability of these models to achieve high sensitivity and precision on the normal class. This ensures that the FPs are minimized not only for the covid but also for the pneumonia class and can potentially help alleviate the burden on the healthcare system.

Regarding NASNetLarge, a precision of 100% for the covid class is observed which means that there were no normal or pneumonia classes falsely missclassified as covid. Furthermore the model would accurately identify “COVID-19 negative” cases 100% of the time but presents a low sensitivity value. Confirmed COVID-19 cases would be able to be identified almost only half the time. Additionally, the model presents a moderate value of 73% for F1-score in covid class. Indeed, this low value is justified by a large number of FNs. Figure 4(c) depicts that the covid class has 11 missclassified cases in total as normal or pneumonia. As expected, this is not acceptable when dealing with such a contagious virus.

Although MobileNetV2 and DenseNet201 present the worst results in terms of accuracy, they outperform VGG16 in terms of specificity and precision (100% for both metrics) for covid class. However, as the results suggest, the sensitivity is one of the most important metric in the particular disease. The extremely low value of 12% depicted for both models can have devastating effects regarding virus spread. Only 12% of confirmed COVID-19 cases would accurately identified correctly. Furthermore, the low value of 21% concerning F1-score in covid class implies many FNs. Indeed, both models presents 23 FNs as illustrated in Figures 4(d), 4(e). A real-life interpretation of a False Negative instance is the erroneous assumption that the patient is “COVID-19 negative” with what this entails in relation to the spread of the virus and public health.

Furthermore, we visualized the loss and the accuracy of the same CNNs during their training in Figure 2. Specifically, Figures 2(a), 2(b), 2(c), 2(d) and 2(e) demonstrate the training/validation loss/accuracy of VGG16, VGG19, NASNetLarge, MobileNetV2 and DenseNet201, respectively. The two best models (Figure 2(a) and 2(b)), demonstrate a smooth training process during which the loss gradually decreases and the accuracy increases. Moreover, it can be observed that the accuracy of both training and validation do not deviate much from one another in most cases, a phenomenon that can also be observed for the training and validation loss, indicating that the models do not overfit. On the other hand, the rest of the models not only present a low accuracy, but their validation loss is either increasing or fluctuating. In the case of NASNetLarge (Figure 2(c)), which presents a relatively high accuracy (in the range of 75%), the fluctuating validation loss means that the model most probably overfits. Another interesting fact is that the models with the least number of layers (VGG16 and VGG19) achieve a better classification performance. This can be explained by the fact that neural networks with more hidden layers require more training data, thus an even larger number of X-Ray samples needs to be provided in these networks.

**Figure 2:**
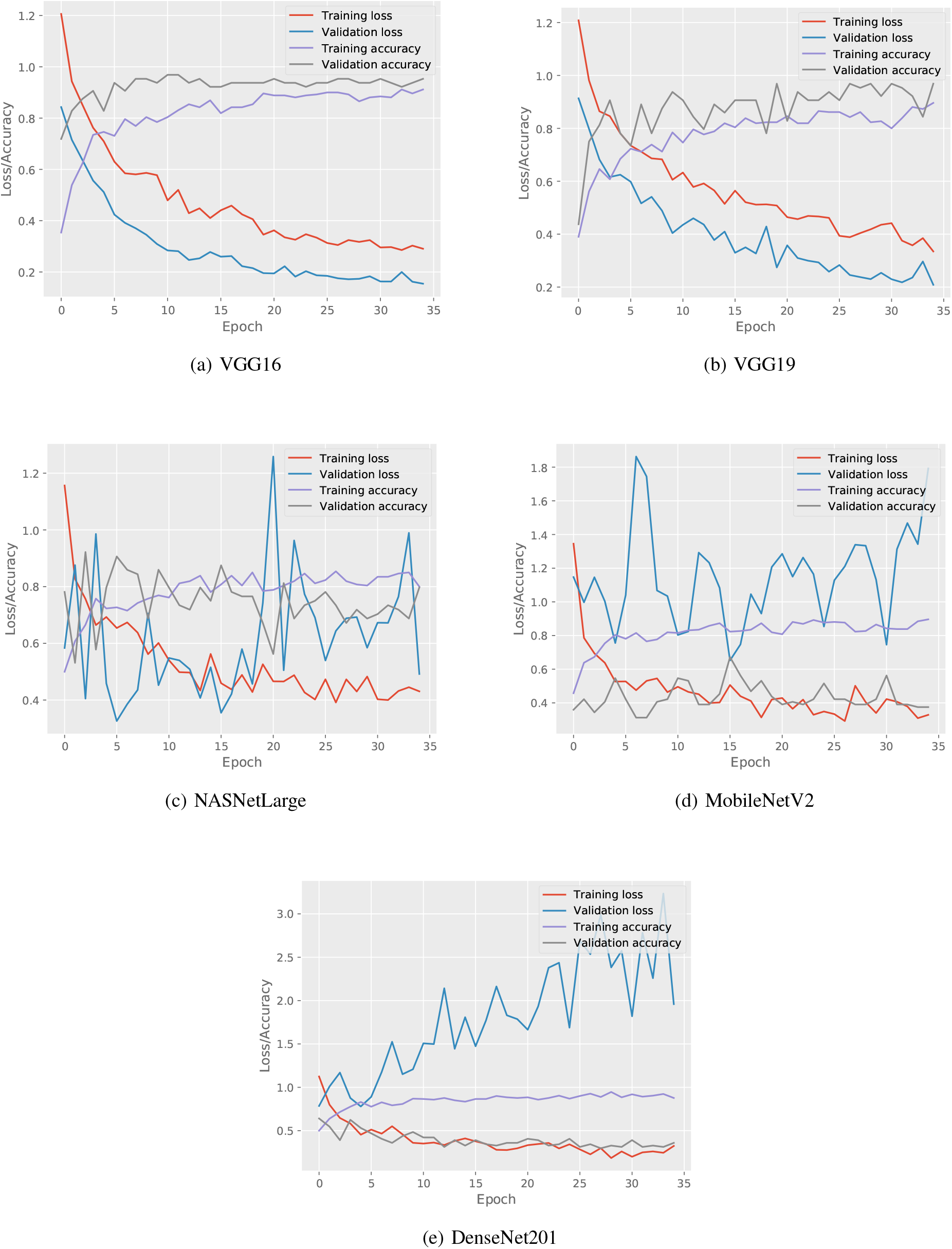
Accuracy and loss (train and test) of deep learning models.

As illustrated in Table 1 all the CNN models present quite high precision and specificity values in covid class even if the overall accuracy is low, except for VGG16 and VGG19. Confusion matrices confirm this as the false positives are zero (only 1 FP in VGG16) as illustrated in Figure 4. However, the sensitivity is extremely low in some models. For example, InceptionV3 and DenseNet201 present the lowest values of sensitivity with 4% and 12% respectively. This practically means, the models are not able to detect the confirmed COVID-19 cases which is likely to cause disastrous results. Furthermore, one important observation is that sensitivity presents high values for pneumonia class except in InceptionResNetV2 model. This ensures that patients with common bacterial pneumonia will not missclassified as covid.

Figure 3 depicts the execution time (in seconds) of each CNN. The largest execution times are presented for the most accurate models. Specifically, NasNetLarge exhibits the highest execution time followed by VGG19 and VGG16. This can be explained by the fact that these models consist of the largest number of parameters. MobileNetV2 presents the lowest execution time and is included along with DenseNet201 in the models with the worst overall accuracy. Nevertheless, InceptionV3, Xception and InceptionResNetV2 present smaller execution times even if the accuracy is much better than DenseNet201.

**Figure 3:**
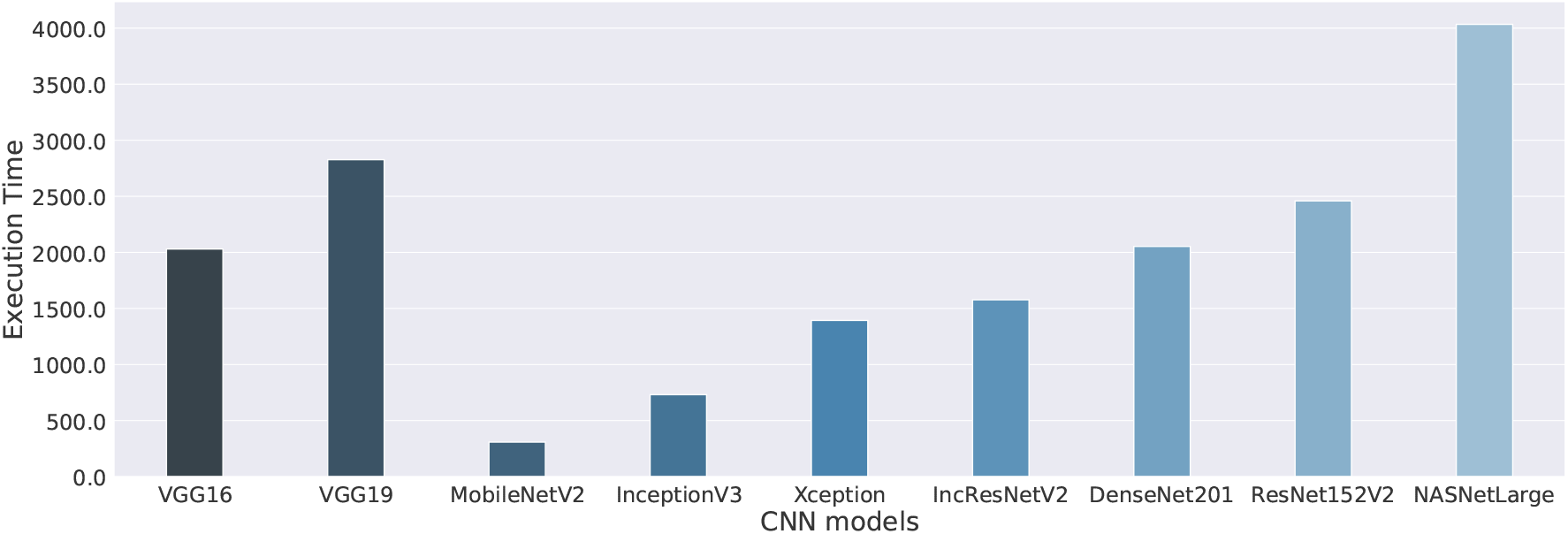
Execution time of all deep learning models on CPU.

**Figure 4:**
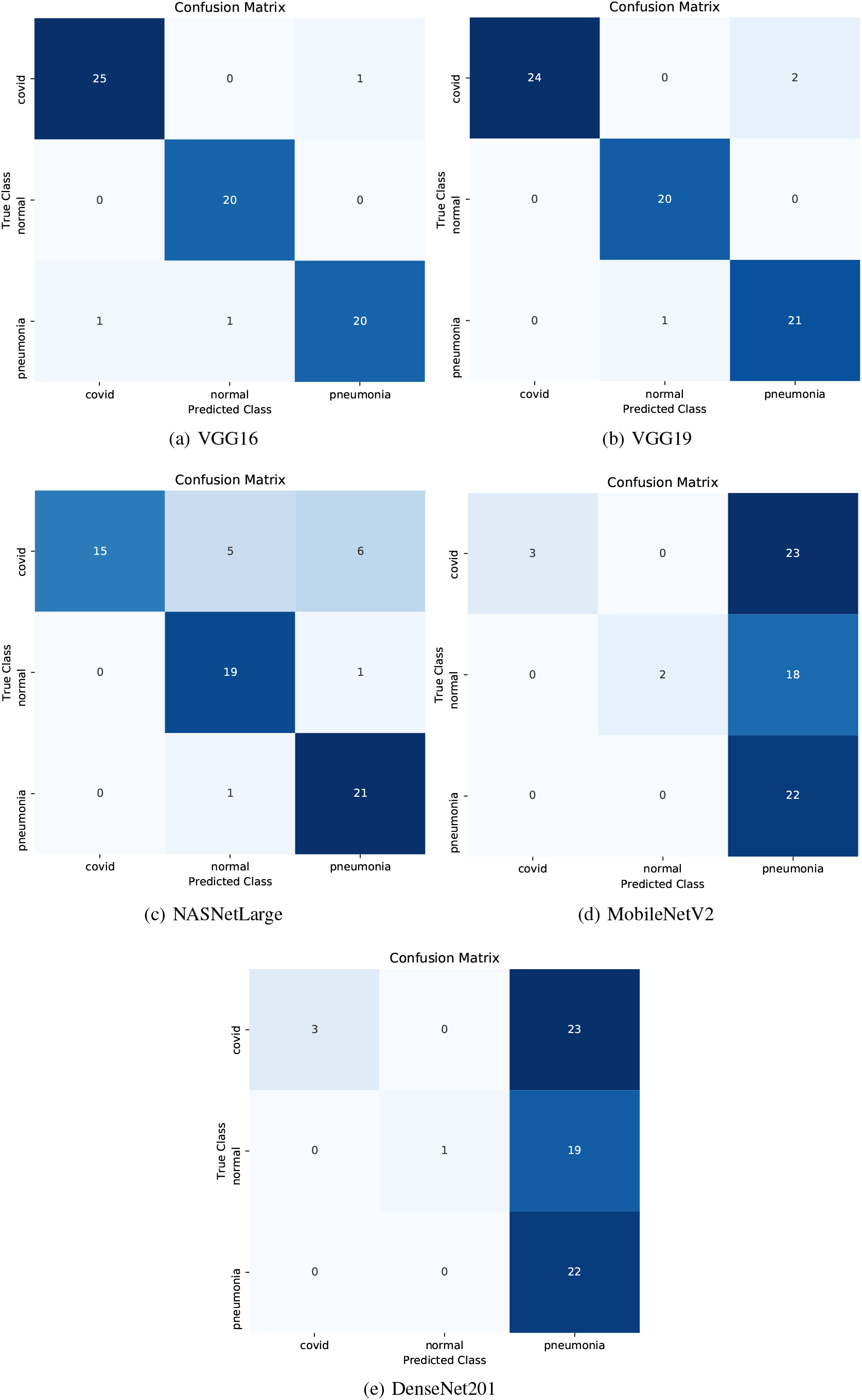
Confusion matrix of deep learning models.

## 5 Conclusion and Future Work

In this work, a study was conducted and presented for the detection of patients positive to COVID-19, a pandemic that infected a large amount of the human population in the first semester of the year 2020. Specifically, the study presented and employed 9 well-known Convolutional Neural Networks (CNNs) for the classification of X-Ray images originating from patients with COVID-19, pneumonia and healthy individuals. Research findings indicated that CNNs have the potential to detect respiratory diseases with high accuracy, although a large amount of sample images needs to be collected. Specifically, VGG16 and VGG19 achieve an overall accuracy of 95%. The high values associated with sensitivity, specificity and precision of covid class, imply the ability of these models to detect positive and/or negative COVID-19 cases accurately thus reducing as much as possible the virus spread to the community. As the results show, determining the most effective model for this classification task involves several performance metrics. Furthermore, one of the very encouraging results is the ability of the aforementioned CNNs to achieve high sensitivity and precision on the normal class thus ensuring the minimization of false positives regarding infection classes which can potentially help alleviate the burden on the healthcare system. Finally, we would like to emphasize that these methods should not be used directly without clinical diagnosis. For future work, we intend to train the CNNs on more data and to evaluate more architectures for the case of COVID-19 detection.

## Data Availability

The dataset used in this research work is available publicly on Github

https://github.com/AntonisMakris/COVID19-XRay-Dataset

## Footnotes

2 Chest X-Ray Images (Pneumonia), https://www.kaggle.com/paultimothymooney/chest-xray-pneumonia

3 https://github.com/AntonisMakris/COVID19-XRay-Dataset

